# Prenatal alcohol exposure and white matter microstructural changes across the first 6-7 years of life: A longitudinal diffusion tensor imaging study of a South African birth cohort

**DOI:** 10.1101/2023.06.30.23292077

**Authors:** KA Donald, CJ Hendrikse, A Roos, CJ Wedderburn, S Subramoney, JE Ringshaw, L Bradford, N Hoffman, T Burd, KL Narr, RP Woods, HJ Zar, SH Joshi, DJ Stein

## Abstract

Prenatal alcohol exposure (PAE) can affect brain development in early life, but few studies have investigated the effects of PAE on *trajectories* of white matter tract maturation in young children. Here we used diffusion weighted imaging (DWI) repeated over three time points in a South African birth cohort, to measure the effects of PAE on patterns of white matter microstructural development during the pre-school years. A total of 342 scans acquired from 237 children as neonates (N=82 scans: 30 PAE; 52 controls) and at ages 2-3 (N=121 scans: 27 PAE; 94 controls) and 6-7 years (N=139 scans: 45 PAE; 94 controls) were included. Maternal alcohol use during pregnancy and other antenatal covariates were collected from 28-32 weeks’ gestation. Linear mixed effects models were implemented to investigate the effects of PAE on fractional anisotropy (FA) and mean diffusivity (MD) in specific white matter tracts over time, while adjusting for child sex and maternal education. We found significant PAE-by-time effects on trajectories of FA development in the left superior cerebellar peduncle (SCP-L: p=0.001; survived FDR correction) and right superior longitudinal fasciculus (SLF-R: p=0.046), suggesting altered white matter development among children with PAE. Compared with controls, children with PAE demonstrated a more rapid change in FA in these tracts from the neonatal period to 2-3 years of age, followed by a more tapered trajectory for the period from 2-3 to 6-7 years of age, with these trajectories differing from unexposed control children. Given their supporting roles in various aspects of neurocognitive functioning (i.e., motor regulation, learning, memory, language), altered patterns of maturation in the SCP and SLF may contribute to a spectrum of physical, social, emotional, and cognitive difficulties often experienced by children with PAE. This study highlights the value of repeated early imaging in longitudinal studies of PAE, and focus for early childhood as a critical window of potential susceptibility as well as an opportunity for early intervention.

## 1. INTRODUCTION

Healthy brain development involves a delicate pattern of complex neurobiological processes that begin early in fetal development and continue throughout childhood (Gilmore et al., 2018). Maternal alcohol use during pregnancy may have teratogenic effects, potentially leading to altered trajectories of child structural and functional neurodevelopment and a wide spectrum of negative neurophysiological outcomes (known as fetal alcohol spectrum disorder [FASD]; Donald et al., 2015; Lebel et al., 2021). Consequently, children with PAE may experience significant long-term difficulties in domains of cognitive, behavioural, and social-emotional functioning (Kar et al., 2021). Investigating the longitudinal effects of PAE on neurodevelopmental trajectories in early childhood may help identify mechanisms that can be targeted for intervention.

Magnetic resonance imaging (MRI) studies, especially those utilizing diffusion weighted imaging (DWI), have significantly broadened our understanding of typical white matter macro- and microstructural development across the lifespan (Ghazi Sherbaf et al., 2019; Lebel & Deoni, 2018). White matter comprises about 50% of brain tissue and consists of billions of nerve cell axons which carry electrical impulses, allowing communication between distant and neighbouring brain regions. The myelin sheaths that surround neuronal axons allow for faster and more efficient transmission of electrical impulses. Although all major white matter tracts begin to develop before birth, myelination occurs predominantly and most rapidly in the first two years of postnatal life and continues at a slower pace up to about 30 years of age (Lebel & Deoni, 2018). DWI metrics—including fractional anisotropy (FA) and mean diffusivity (MD)—are sensitive to the microstructural properties of white matter (e.g., myelination). FA, the most widely reported DWI metric, typically increases with brain development, presumably reflecting increased organization of white matter tracts, including myelination (Gilmore et al., 2018; Lebel et al., 2019). Lower FA is often considered to represent reduced tract microstructural integrity, although the interpretation of relative value of direction may be less clear for the first years of life (Barnea-Goraly et al., 2005).

PAE may affect white matter development by altering processes of glial cell proliferation and migration in utero, thereby disrupting the development of myelin sheaths around neuronal axons after birth (Goodlett et al., 2005; Wilhelm & Guizzetti, 2016). Cross-sectional neuroimaging studies have demonstrated the potential effects of PAE on white matter microstructure in association, projection, callosal and brain stem tracts across various age-groups, including infants, children, adolescents, and adults (Ghazi Sherbaf et al., 2019; Kar et al., 2022). One of the most widely reported findings is reduced integrity or FA of the corpus callosum (CC), the largest commissural tract connecting the left and right hemispheres of the brain (Ghazi Sherbaf et al., 2019). Altered CC integrity has been associated with poorer cognitive performance, including information processing and eyeblink conditioning in some studies (Fan et al., 2016); however results have been mixed (refs). More widespread microstructural changes following PAE have also been reported, specifically of the cerebellar peduncles (Donald et al., 2015; Roos et al., 2021; Fan et al., 2016) and cingulum (Lebel et al., 2008; Paolozza et al., 2017; Santhanam et al., 2011), corticospinal tract, uncinate fasciculus as well as both the inferior and superior longitudinal fasciculi (Donald et al., 2015; Roos et al., 2021).

Due to a lack of longitudinal investigations (Lebel et al., 2021), as well as a paucity of studies in young children specifically (Ghazi Sherbaf et al., 2019; Kar et al., 2022), our understanding of the effects of PAE on early life trajectories of white matter microstructure remains limited. Despite early neuroplasticity, little is known about the extent to which certain white matter microstructural changes detected in infants and very young children with PAE may dissipate or intensify towards middle-childhood. To address these gaps in the literature, we aimed to investigate the effects of PAE on the trajectories of white matter microstructural maturation (using DWI metrics FA and MD) in a cohort of children living in a peri-urban Western Cape community in South Africa, followed from birth to age at school entry (i.e., 6-7 years). In this cohort, which forms part of the Drakenstein Child Health Study (DCHS), we have previously found altered microstructure of the superior longitudinal fasciculus in infants with PAE, with changes in the inferior cerebellar peduncles associated with abnormal neonatal neurobehaviour at the same age (Donald et al., 2015). In a follow-up study at age 2-3 years, we observed microstructural alterations in these same regions, and additionally in the uncinate fasciculus, corticospinal tract, fornix stria terminalis, and sagittal stratum (Roos et al., 2021). Repeated scanning over these early years allows detection of PAE-related changes in white matter microstructure over time, and may elucidate windows of maximal impact of PAE on early trajectories of white matter development.

## 2. METHODS

### 2.1. Study design and participants

Participants were part of the Drakenstein Child Health Study (DCHS), an ongoing population-based birth cohort study investigating early determinants of child health and development, conducted in the peri-urban Drakenstein region of the Western Cape, South Africa. The Drakenstein community is comprised of about 200 000 individuals of low socio-economic status living in informal housing, and rates of unemployment are high. More than 90% of women in this region rely on the public health sector for primary health care, including antenatal care and child health services (Stein et al., 2015; Zar et al., 2015). The rate of substance misuse is high in this community, reflecting the broader South African population more generally. Indeed, South Africa has the highest reported prevalence of FASD in the world, with rates as high as 310 per 1000 children living in rural communities (May et al., 2022).

The procedures of the DCHS have been described in detail elsewhere (Stein et al., 2015; Zar et al., 2015). Briefly, pregnant women aged 18 years or older were recruited at 20-28 weeks’ gestation while attending routine antenatal care at one of two primary healthcare clinics in the Drakenstein area. Mothers provided written informed consent for themselves and their children to participate in the study, and background data were collected (i.e., socio-demographics, psychosocial risk factors). Ethics approval for the main DCHS and this brain imaging sub-study were obtained from the University of Cape Town Health Sciences Faculty, Human Research Ethics Committee (401/2009, 525/2012, 044/2017) and the Western Cape Department of Provincial Health Research Committee (2011RP45).

A subset of children with and without PAE from mothers participating in the DCHS were included in the current brain imaging sub-study and scanned at three time points over the first 6-7 years of life. Exclusion criteria for the children were: (i) the mother had a positive urine toxicology result during pregnancy for other drugs of abuse (i.e., besides alcohol and tobacco), (ii) low Apgar score (<7 at 5 min), (iii) admittance to the neonatal intensive care unit, (iv) an identified genetic syndrome or congenital abnormalities, and/or (v) standard MRI contraindications (Donald et al., 2015; Wedderburn et al., 2020). Supplementary Figure 1 presents a breakdown of participant dropout and exclusion rates for the three time points. A total of 342 scans acquired on 237 children (67 PAE, 170 controls) across the three time points were included in the current analysis comprising 82 neonates (30 PAE, 52 controls), 121 2-3-year-olds (27 PAE, 94 controls), and 139 6-7-year-olds (45 PAE, 94 controls). Figure 1 depicts the age and time between scans for all included children by PAE status. Fifty-nine percent of the children (N=140: 34 PAE, 106 controls) were scanned at one time point, while 41 percent (N=97: 33 PAE, 64 controls) were scanned at two or three time points (Figure 1).

**Figure 1:**
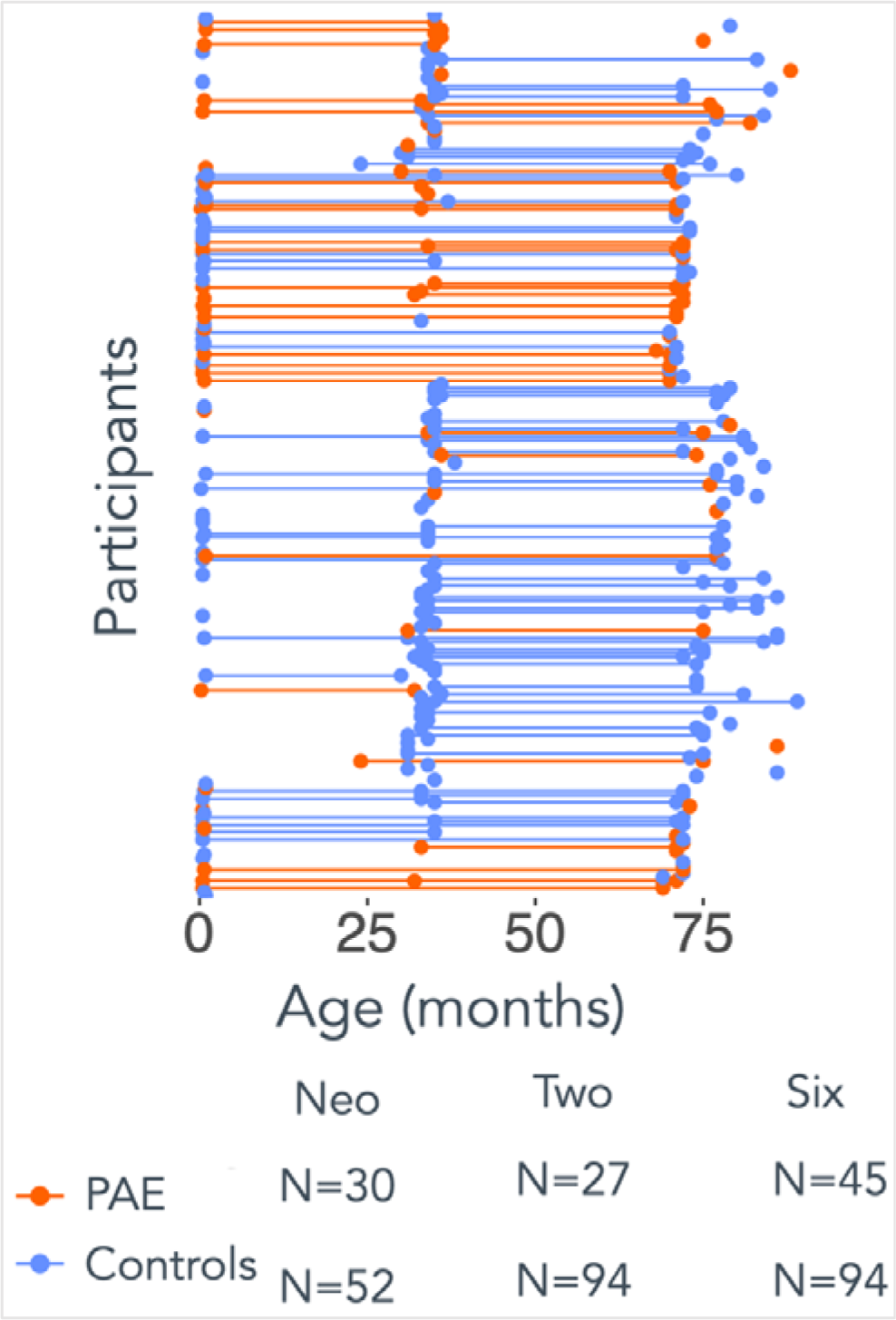
Age and time between scans by prenatal alcohol exposure (PAE) status. Each dot represents a scan (N=342), connecting lines represent the time between scans, and colour indicates the PAE status for each participant.

### 2.2. Maternal assessments

All mothers completed the World Health Organization’s Alcohol, Smoking and Substance Involvement Screening Test (ASSIST) between 28-32 weeks’ gestation to assess comorbid substance use (Humeniuk et al., 2008); this self-report questionnaire has been validated for use in South Africa (Jackson et al., 2010). The children born to mothers with a total ASSIST score above 11 on the alcohol questions, and/or who indicated moderate-severe alcohol use during pregnancy (i.e., >2 drinks twice per week in any trimester), were categorized as alcohol exposed (i.e., PAE group). Maternal urine cotinine levels were additionally measured to determine active tobacco smoking status during pregnancy (i.e., cotinine level >500 ng/mL). Mothers also completed the Beck Depression Inventory during pregnancy, and the Edinburgh Postnatal Depression Scale at six months postpartum, to measure prenatal and postnatal depressive symptomatology (Beck et al., 1988; Cox et al., 1987; Tsai et al., 2013).

### 2.3. Child brain imaging

Imaging protocols were tailored for each of the three time points to be optimal for the children’s developmental stage (Roos et al., 2022). Table 1 describes the scanner specifications, imaging modalities, and acquisition parameters relevant to each time point. Two DWI sets were obtained at each time point, in the transverse plane with both anterior– posterior and posterior–anterior phase encoding to control for anatomic distortions and increase signal-to-noise. T2-weighted (time point 1) or T1-weighted (time points 2 and 3) structural MRI (sMRI) scans were also collected and provided an individual anatomical reference volume for each child during image processing. Anthropometric data (i.e., weight, height, head circumference) were collected at each time point, and health information at birth were extracted from hospital records. Children were asleep during scanning at the first two time points, but awake and quiet during the last.

**Table 1:**
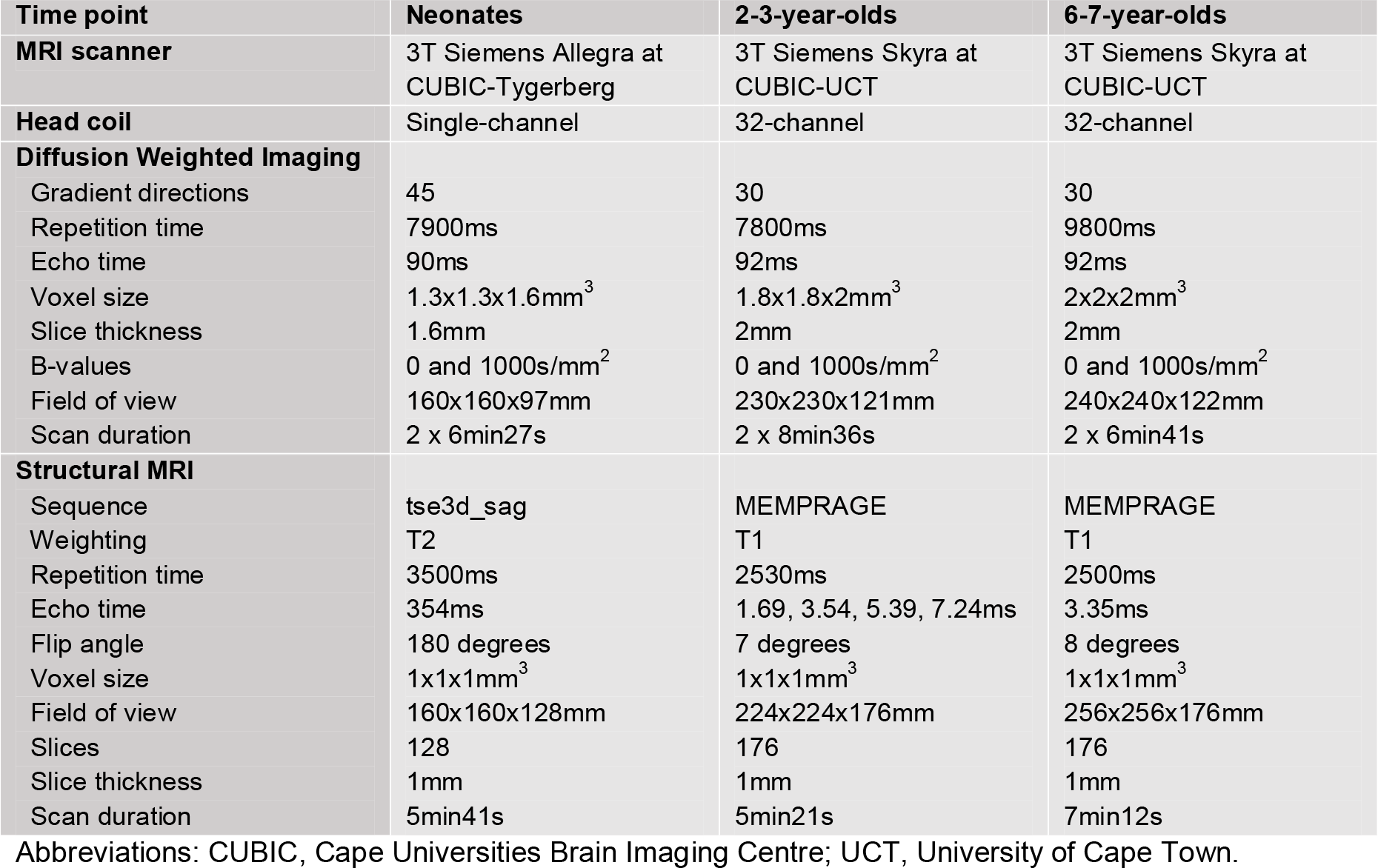
Imaging protocol for each time point.

| Time point | Neonates | 2-3-year-olds | 6-7-year-olds |
| --- | --- | --- | --- |
| **MRI scanner** | 3T Siemens Allegra at CUBIC-Tygerberg | 3T Siemens Skyra at CUBIC-UCT | 3T Siemens Skyra at CUBIC-UCT |
| **Head coil** | Single-channel | 32-channel | 32-channel |
| **Diffusion Weighted Imaging** |  |  |  |
| Gradient directions | 45 | 30 | 30 |
| Repetition time | 7900ms | 7800ms | 9800ms |
| Echo time | 90ms | 92ms | 92ms |
| Voxel size | 1.3x1.3x1.6mm<sup>3</sup> | 1.8x1.8x2mm<sup>3</sup> | 2x2x2mm<sup>3</sup> |
| Slice thickness | 1.6mm | 2mm | 2mm |
| B-values | 0 and 1000s/mm<sup>2</sup> | 0 and 1000s/mm<sup>2</sup> | 0 and 1000s/mm<sup>2</sup> |
| Field of view | 160x160x97mm | 230x230x121mm | 240x240x122mm |
| Scan duration | 2 x 6min27s | 2 x 8min36s | 2 x 6min41s |
| **Structural MRI** |  |  |  |
| Sequence | tse3d_sag | MEMPRAGE | MEMPRAGE |
| Weighting | T2 | T1 | T1 |
| Repetition time | 3500ms | 2530ms | 2500ms |
| Echo time | 354ms | 1.69, 3.54, 5.39, 7.24ms | 3.35ms |
| Flip angle | 180 degrees | 7 degrees | 8 degrees |
| Voxel size | 1x1x1mm<sup>3</sup> | 1x1x1mm<sup>3</sup> | 1x1x1mm<sup>3</sup> |
| Field of view | 160x160x128mm | 224x224x176mm | 256x256x176mm |
| Slices | 128 | 176 | 176 |
| Slice thickness | 1mm | 1mm | 1mm |
| Scan duration | 5min41s | 5min21s | 7min12s |

Abbreviations: CUBIC, Cape Universities Brain Imaging Centre; UCT, University of Cape Town.

### 2.4. Image processing

All MRI data were quality checked before preprocessing. Participants had to have at least one structural MRI scan and one DWI scan in each phase-encoding direction without disruptive movement or other artifacts for inclusion. DWI scans were preprocessed on the Centre for High Performance Computing cluster (CHPC, Cape Town) using TORTOISE software (Irfanoglu et al., 2018; Pierpaoli et al., 2010), which was chosen due to its superior correction and registration ability in pediatric samples prone to movement during scanning (Roos et al., 2021; Taylor et al., 2016). In the neonates, T2-weighted sMRI images were registered to the University of North Carolina (UNC) neonate structural template (Shi et al., 2011) given the relatively immature state of tract formation compared to the other time points (Roos et al., 2022). The registered image was then used as an anatomical reference image in TORTOISE. In the other time points, T1-weighted sMRI images were inverted to have similar contrast to the diffusion b0 volume, and used as an anatomical reference volume in TORTOISE. The DIFF PREP and DR BUDDI modules of TORTOISE were used, respectively, to (i) compute motion, eddy currents, and echo-planar imaging (EPI) distortion corrections, and (ii) merge encoded sets and perform further EPI distortion corrections.

After preprocessing, the Tract-based Spatial Statistics (TBSS) pipeline of FMRIB Software Library (FSL) was used to perform diffusion tensor parameter fitting and to extract diffusion parameters (Smith et al., 2006). Preprocessed FA images were first created by applying brain extraction using BET and tensor extraction using DTIFIT (Smith, 2002). Thereafter, the 4 steps of TBSS were applied as described in Roos et al. (2021). Importantly, since we had a pediatric sample for whom the use of an adult MNI template would be inappropriate, we used study-specific templates as the registration target in the second step of TBSS within each time point. The use of a study-specific template significantly enhances registration quality (Bach et al., 2014; Smith et al., 2006). Finally, the mean FA skeleton was used to map and derive MD images, and summary metrics were extracted for 48 white matter tracts using the Johns Hopkins University ICBM-DTI-81 atlas (Mori et al., 2008).

### 2.5. Statistical analysis

We conducted all statistical analyses in R version 4.2.1. Linear mixed effects models were fitted with restricted maxium likelihood (REML) to accommodate missing data, and implemented using lme4 (Bates et al., 2015) and lmerTest (Kuznetsova et al., 2017). We tested the effects of PAE, time, and the interaction of PAE with time as predictors while controlling for sex and maternal education as regressors on FA and MD of those salient white matter tracts implicated by PAE in our prior work. Figure 2 illustrates the investigated tracts, which included the left and right superior longitudinal (SLF-L/R) and uncinate fasciculus (UF-L/R), left and right sagittal stratum (SS-L/R), left and right inferior (ICP-L/R) and superior cerebellar peduncles (SCP-L/R), corpus callosum genu (CC-G) and splenium (CC-S), fornix, and left and right superior corona radiata (SCR-L/R). The participant was included as a random effect. Specifically, the fixed effects model was given by FA ∼ PAE status + Time + Sex + Maternal education + (PAE status x Time). We tested the omnibus model with all three time points together followed by pairwise comparisons between two time points, separately for neonates and 2-3-year-old children, neonates and 6-7-year-old children, and 2-3-year-old and 6-7-year-old children. Post-hoc cross-sectional analysis for group differences between children with PAE and controls within each time point was performed with ANOVA after adjusting for sex and maternal education. Additionally, post-hoc sensitivity analyses were performed to test the robustness of our findings while including prenatal tobacco exposure as an additional covariate, given its known association with white matter microstructure in this cohort (Roos et al., 2021). We used the Benjamini Hochberg false discovery rate (FDR) to correct for multiple comparisons within each diffusion measure across all tracts, though we report findings that survived correction and those that did not, given the strong a priori inclusion of these tracts based on findings in our own cohort.

**Figure 2:**
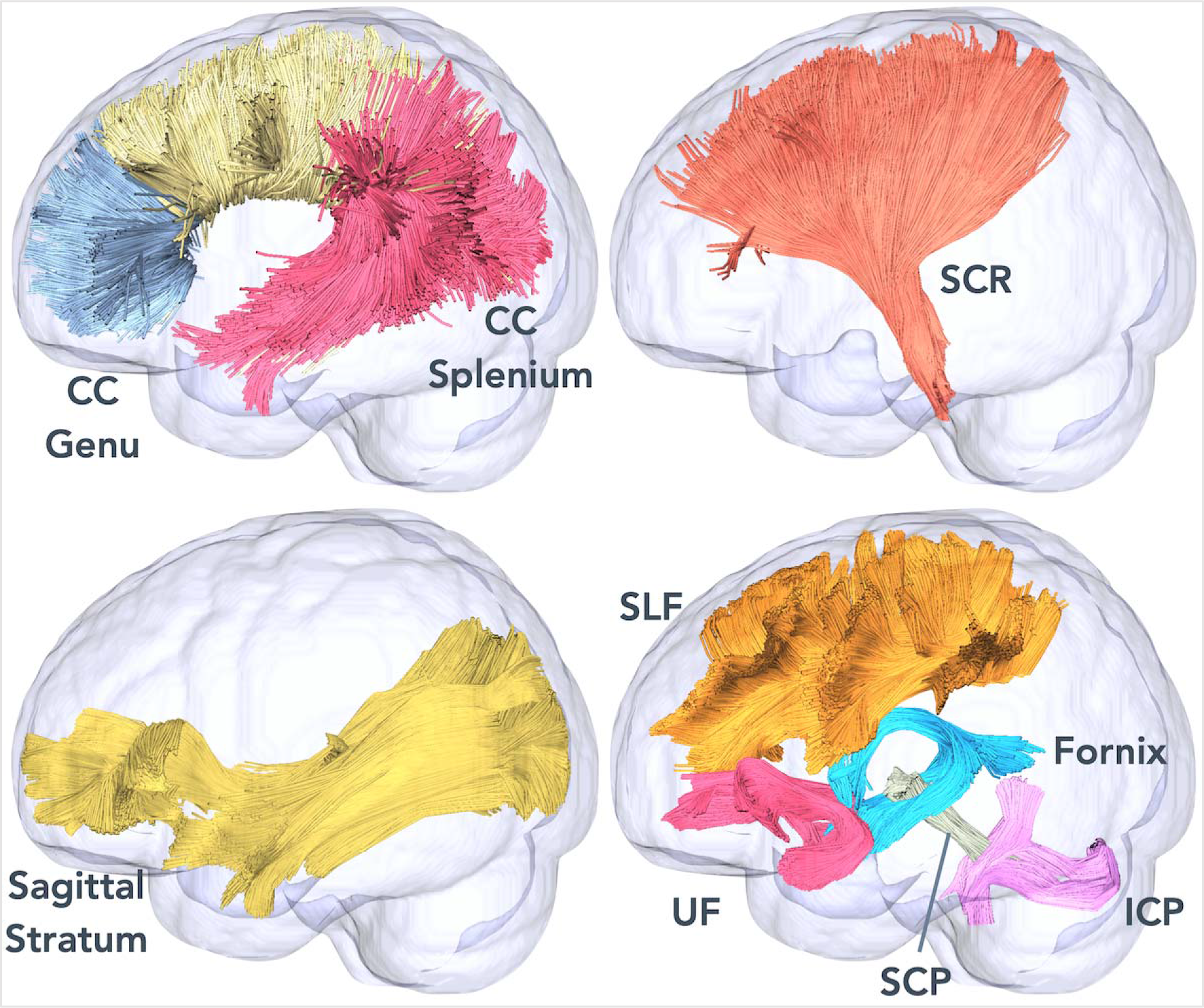
Illustration of investigated tracts on glass brains. CC=corpus callosum, SCR=superior corona radiata, SLF=superior longitudinal fasciculus, UF=uncinate fasciculus, SCP=superior cerebellar peduncle, ICP=inferior cerebellar peduncle.

## 3. RESULTS

### 3.1. Socio-demographic and psychosocial group differences

Table 2 outlines the socio-demographic and psychosocial characteristics of PAE and control groups. PAE and control groups were similar in terms of sex and gestational age at delivery within each time point. A 4-month difference in mean age was present at the 6-7-year time point, but not at the neonatal and 2-3 year points, with PAE children being slightly younger than controls at scanning. However, the age range of the PAE and control groups were similar (68-88 and 69-89 months respectively). Similarly, children with PAE had smaller anthropometric measurements in terms of height, weight, and head circumference than controls at the 6-7 year time point only. Maternal tobacco use during pregnancy was higher in PAE children compared to controls for all three time points, however we tested the influence of PTE on our main findings in a post-hoc sensitivity analysis. Maternal characteristics were generally matched for children with PAE and controls, in terms of age at enrolment (except for time point 3), educational attainment, employment, monthly income, marital status, household size, unplanned pregnancy, HIV infection, and pre- and postnatal depression.

**Table 2:**
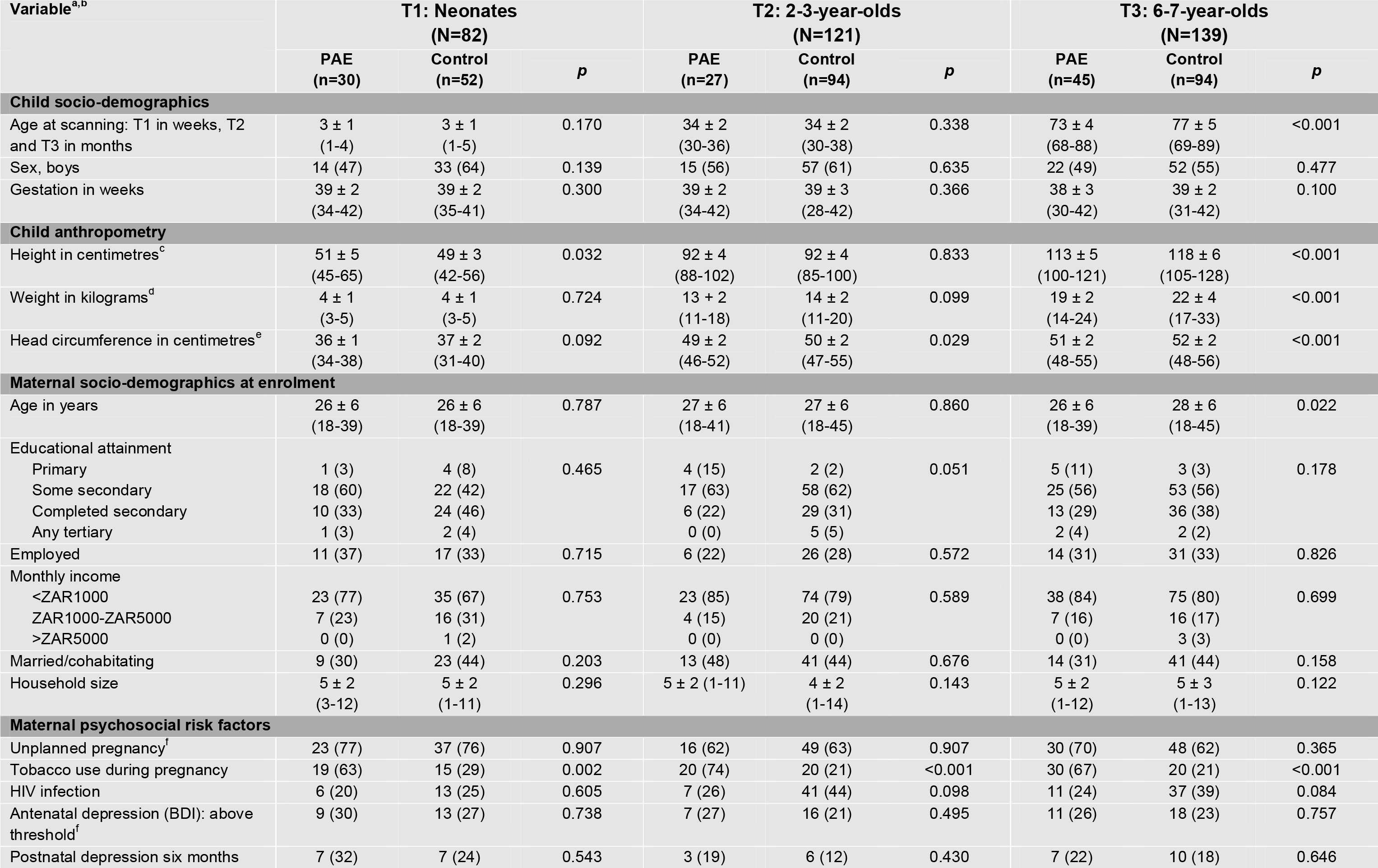
Socio-demographic characteristics and psychosocial risk factors of children and mothers across the three time points.

### 3.2. PAE effects on FA and MD across development

We found significant main effects of PAE on FA and MD across the period from birth to 6-7 years (see Table 3 for a summary of results or Supplementary Table 1 for full results). Children with PAE demonstrated greater FA in the SCP-L/R (p=0.021; Figure 3), ICP-R (p=0.011), SS-L (p=0.001), and SLF-L (p=0.025; Figure 3), and reduced MD in the SLF-R (p=0.031; Figure 4), compared with controls, though these findings did not survive FDR corection. Starting with the neonates, the differences in FA between children with PAE and controls were minimal; however, FA in the 2-3-year-old children with PAE was higher than that of the controls. These differences decreased again as both the groups reached 6-7 years of age.

**Figure 3:**
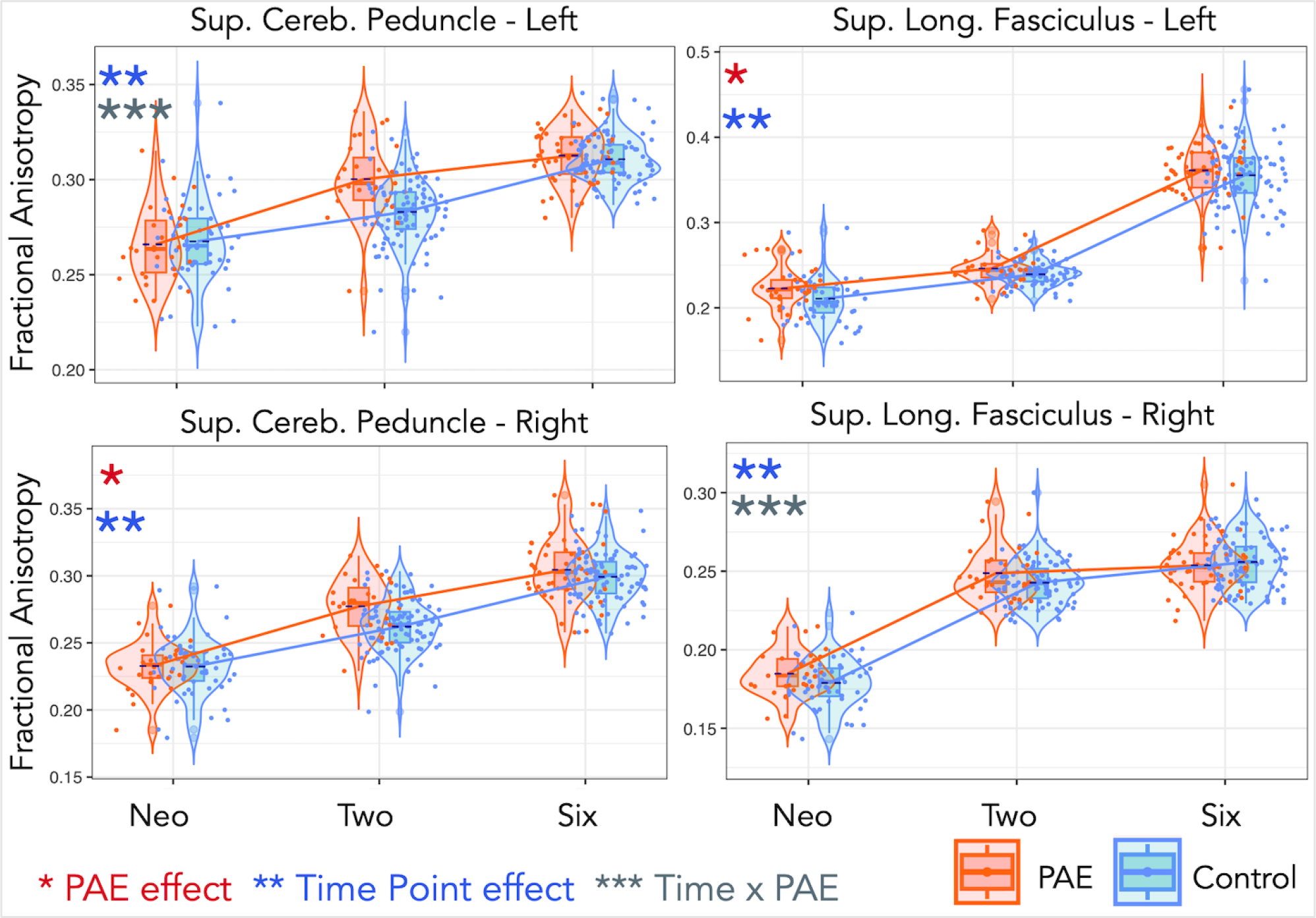
Trajectories of fractional anisotropy (FA) for the left and right tracts of the superior cerebellar peduncle and the superior longitudinal fasciculus for both PAE and controls through birth, 2-3-, and 6-7-year time points in development. The SCP-L was significant after correction. One star (*) shows a significant PAE effect, two stars (**) show a time effect, and three stars (***) show a PAE-by-time interaction effect. Jitter is added for visualization purposes (only in the time point direction).

**Figure 4:**
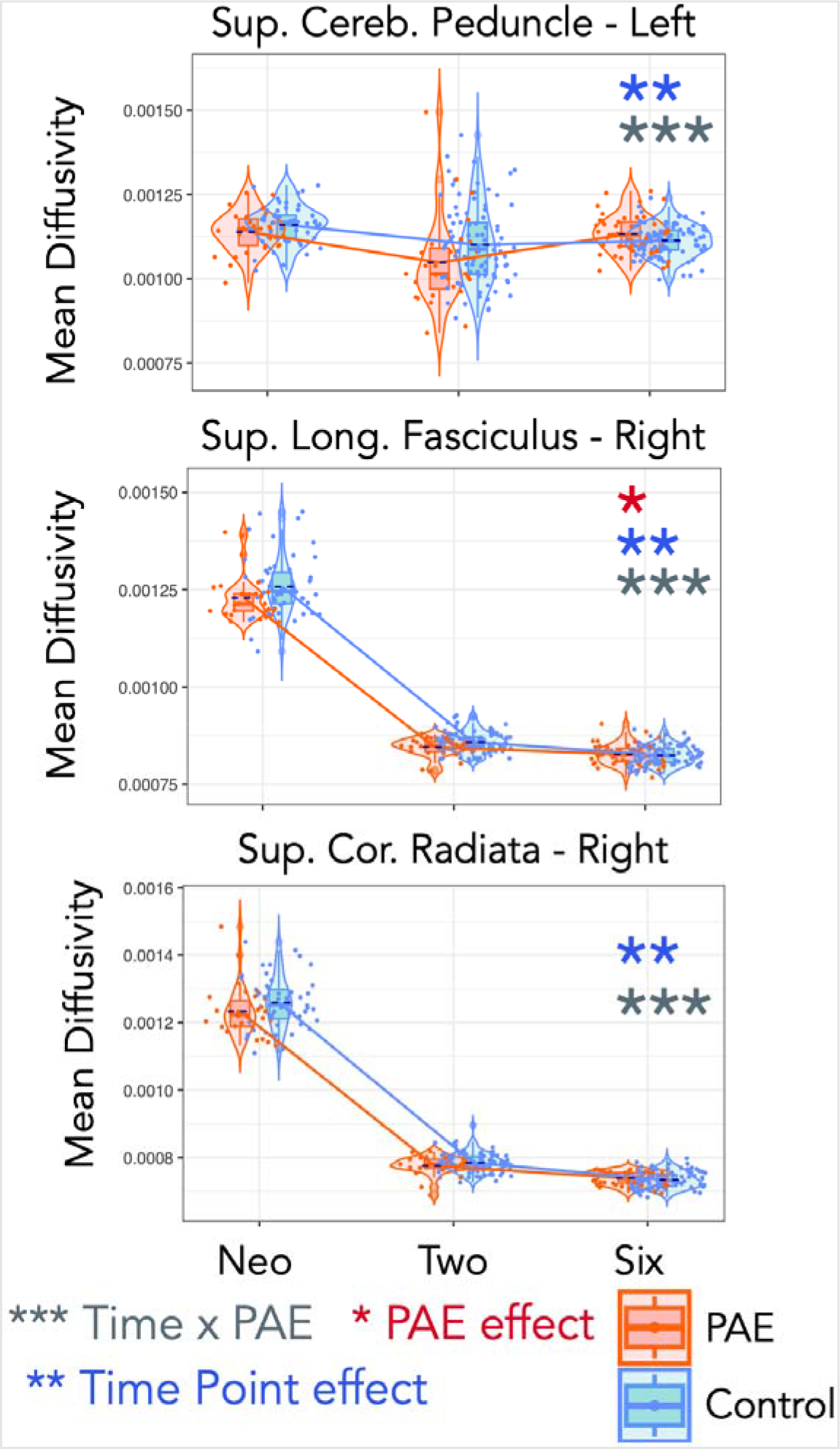
Trajectories of mean diffusivity (MD) for the left superior cerebellar peduncle, and the right superior longitudinal fasciculus and superior corona radiata for both PAE and controls through birth, 2-3-, and 6-7-year time points in development. One star (*) shows a significant PAE effect, two stars (**) show a time effect, and three stars (***) show a PAE-by-time interaction effect (not corrected for multiple comparisons). Jitter is added for visualization purposes (only in the time point direction).

**Table 3:** Longitudinal effects of PAE, Time, and PAE-by-time interaction on FA and MD.

| Tract | L/R | Fractional Anisotropy (FA)<sup>a</sup> |  |  |  |  |  | Mean diffusivity (MD)<sup>a</sup> |  |  |  |  |  |
| --- | --- | --- | --- | --- | --- | --- | --- | --- | --- | --- | --- | --- | --- |
|  |  | PAE |  | Time |  | PAE x Time |  | PAE |  | Time |  | PAE x Time |  |
|  |  | Partial eta<sup>2</sup> | *p* | Partial eta<sup>2</sup> | *p* | Partial eta<sup>2</sup> | *p* | Partial eta<sup>2</sup> | *p* | Partial eta<sup>2</sup> | *p* | Partial eta<sup>2</sup> | *p* |
| Superior longitudinal fasciculus | L | 0.016 | 0.025 | 0.842 | <0.001* | - | ns | - | ns | 0.962 | <0.001* | - | ns |
|  | R | - | ns | 0.864 | <0.001* | 0.027 | 0.046 | 0.016 | 0.031 | 0.951 | <0.001* | 0.022 | 0.038 |
| Sagittal stratum | L | 0.039 | 0.001 | 0.881 | <0.001* | - | ns | - | ns | 0.928 | <0.001* | - | ns |
|  | R | - | ns | 0.897 | <0.001* | - | ns | - | ns | 0.957 | <0.001* | - | ns |
| Uncinate fasciculus | L | - | ns | 0.923 | <0.001* | - | ns | - | ns | 0.935 | <0.001* | - | ns |
|  | R | - | ns | 0.820 | <0.001* | - | ns | - | ns | 0.690 | <0.001* | - | ns |
| Inferior cerebellar peduncle | L | - | ns | 0.563 | <0.001* | - | ns | - | ns | 0.744 | <0.001* | - | ns |
|  | R | 0.026 | 0.011 | 0.512 | <0.001* | - | ns | - | ns | 0.262 | <0.001* | - | ns |
| Superior cerebellar peduncle | L | 0.023 | 0.007 | 0.522 | <0.001* | 0.042 | 0.001* | - | ns | 0.109 | <0.001* | 0.035 | 0.008 |
|  | R | 0.025 | 0.021 | 0.784 | <0.001* | - | ns | - | ns | 0.477 | <0.001* | - | ns |
| Corpus callosum genu | n/a | - | ns | 0.954 | <0.001* | - | ns | - | ns | 0.951 | <0.001* | - | ns |
| Corpus callosum splenium | n/a | - | ns | 0.924 | <0.001* | - | ns | - | ns | 0.978 | <0.001* | - | ns |
| Fornix | n/a | - | ns | 0.248 | <0.001* | - | ns | - | ns | 0.037 | 0.039 | - | ns |
| Superior corona radiata | L | - | ns | 0.926 | <0.001* | - | ns | - | ns | 0.968 | <0.001* | - | ns |
|  | R | - | ns | 0.924 | <0.001* | - | ns | - | ns | 0.965 | <0.001* | 0.021 | 0.047 |

Abbreviations: PAE, prenatal alcohol exposure; L/R, left or right hemisphere; n/a, not applicable; ns, not significant at $p < 0.05$ .
<sup>a</sup> Model adjusting for sex and maternal education.
\* Survived false discovery rate (FDR) correction for multiple comparison.

### 3.3. Time effects on FA and MD across development

As expected, all tracts showed a significant main effect of time (p < 0.0001) for both FA and MD throughout development from the neonatal age to 6-7 years (Table 3, Supplementary Table 1). FA increased with age, while MD decreased with age.

### 3.4. PAE-by-time effects on FA and MD across development

We found a significant PAE-by-time interaction effect for FA in the SCP-L (p=0.001) that survived correction for multiple comparisons, after adjusting for sex and maternal educational attainment. This differential effect in the rate of change of white matter maturation shows a more rapid increase of FA in children with PAE from the neonatal period to 2-3 years of age, and then a more tapered change for the period from 2-3 to 6-7 years of age compared to controls. Conversely, the rate of change in FA was slower in controls in the initial neonatal period up to 2-3 years, and slightly higher from 2-3 to 6-7 years. We also found a PAE-by-time interaction effect for FA in the SLF-R (p=0.046), which did not survive FDR correction. For this tract, FA showed a minimal rate of change for both groups from 2-3 to 6-7 years of age. Figure 3 shows trajectories of FA development across the three time points for the SCP-L/R and SLF-L/R.

We found PAE-by-time interaction effects for MD in the SLF-R (p=0.038), SCP-L (p=0.008), and SCR-R (p=0.047), after adjusting for sex and maternal educational attainment, however, these effects did not survive multiple comparison correction. Figure 4 shows trajectories of MD development across the three time points for these tracts. Complementary to the effects on FA, we observed that MD was lower in children with PAE compared with controls across all time points (except the SCP-L at age 6-7 years) in these tracts.

### 3.5. Post-hoc pairwise comparisons of time points

In a post-hoc analysis, we compared pairwise time points for neonates and 2-3 years, neonates and 6-7 years, and 2-3 and 6-7 years using the same interaction model as defined previously.

#### 3.5.1. Neonates and 2-3-year-olds

We found significant interaction effects between PAE and time for FA in the SCP-L (p=0.004), SCP-R (p=0.039), and UF-R (p=0.032). We also found significant effects of PAE across the neonatal period to 2-3 years for FA in the SCP-L/R (p=0.019, p=0.026), SS-L/R (p=0.011, p=0.037), and SLF-L/R (p=0.007, p=0.015) and for MD in the SCP-L (p=0.039), and SLF-R (p=0.027). These effects did not survive FDR correction (Supplementary Table 2).

#### 3.5.2. Neonates and 6-7-year-olds

There were no significant interaction effects between PAE and time on FA. Significant PAE-by-time effects on MD were evident in the SLF-R (p=0.028), SCP-L (p=0.010), and SCR-R (p=0.033). Moreover, we found nominally significant effects (uncorrected) of PAE for FA in the ICP-R (p=0.033) and SS-L (p=0.021). None of these effects survived multiple comparisons corrections (Supplementary Table 3).

#### 3.5.3. 2-3-year-olds and 6-7-year-olds

We found an interaction effect between PAE and time for FA in the SCP-L (p=0.0007), which was significant after multiple comparisons correction, as shown in Figure 5. The interaction effects for FA in the CC-G (p=0.048), SCR-L (p=0.017), SLF-R (p=0.008), and fornix (p=0.022) did not survive correction. Significant PAE effects (corrected for multiple comparisons) were found for FA in the SCP-L/R (p<0.001, p=0.003), ICP-R (p=0.010), and SS-L (p=0.0008) (Figure 5). We also found significant PAE-by-time effects on MD in the SCP-L (p=0.005), SS-R (p=0.010), and SLF-L (p=0.001), which survived multiple comparisons correction, whereas PAE-by-time effects on MD in the SLF-R (p=0.028), CC-G (p=0.030), and SCR-L (p=0.042) did not survive correction (Supplementary Table 4).

**Figure 5:**
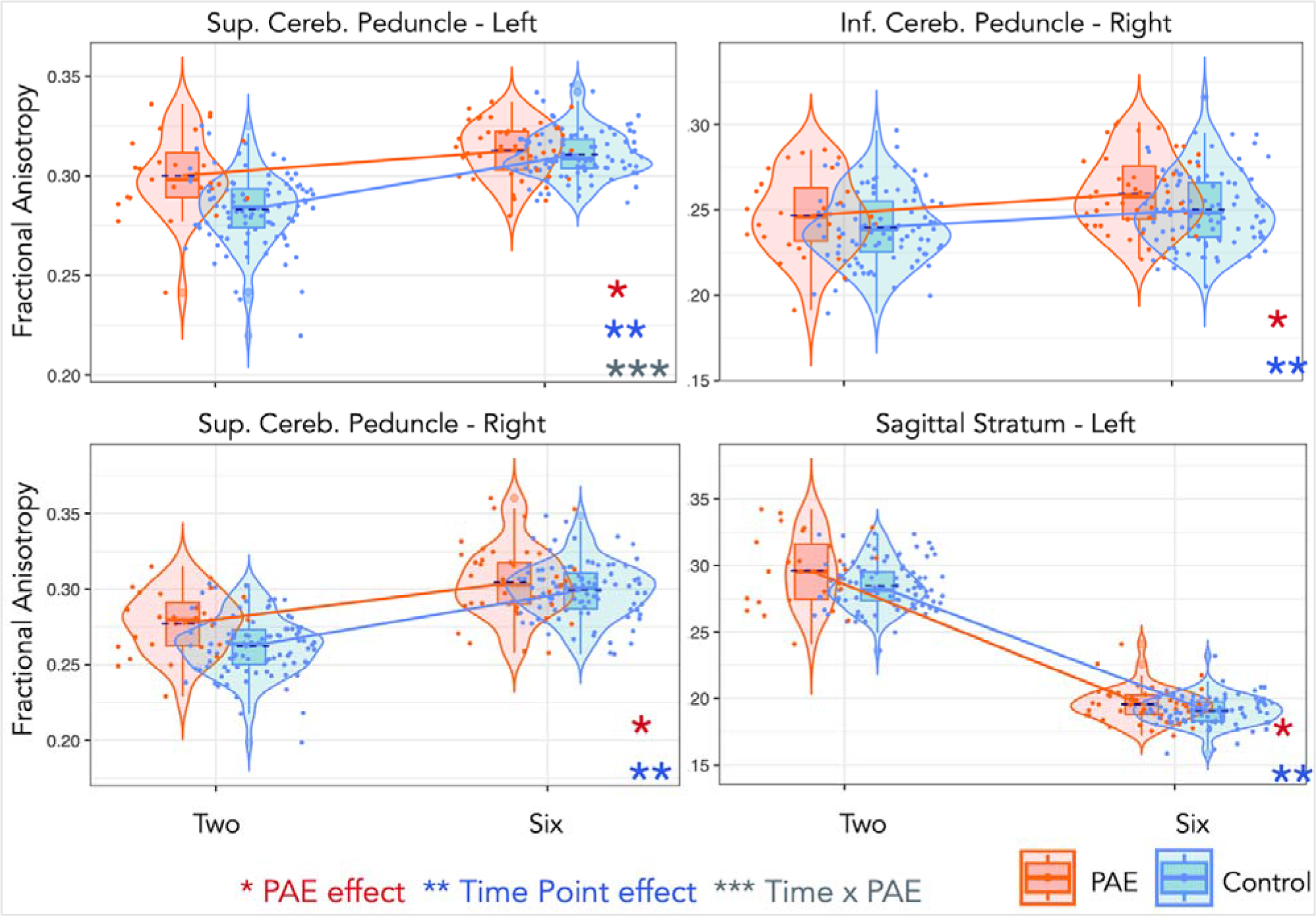
Trajectories of fractional anisotropy (FA) for the left and right tracts of the superior cerebellar peduncle, the left inferior cerebellar peduncle, and the left sagittal stratum for both PAE and unexposed controls through 2-3 and 6-7 years. One star (*) shows a significant PAE effect, two stars (**) show a time effect, and three stars (***) show a PAE-by-time interaction effect. All effects were significant after multiple comparisons correction.

### 3.6. Cross-sectional PAE effects on FA and MD within each time point

Table 4 provides a summary of PAE effects on FA and MD within each time point. At the neonatal age, no significant differences were found in FA or MD in any of the tracts. At the 2-3-year time point, children with PAE demonstrated significantly greater FA than controls in the SCP-L/R (p=0.00015, p=0.0015) when corrected for multiple comparisons. They also had increased FA in the SS-L/R (p=0.017, p=0.048), and SLF-L/R (p=0.038, p=0.041) and decreased MD in the SS-L (p=0.015) and SLF-L/R (p=0.014, p=0.048) compared with controls, however, these differences failed to achieve significance after correction. Lastly, at 6-7 years of age, children with PAE showed higher FA in the ICP-R (p=0.020) and SS-L (p=0.019), and higher MD in the SCP-L (p=0.045) compared with controls, which were not significant after correction for multiple comparisons.

**Table 4:** Summary of cross-sectional PAE effects on FA and MD within each time point.

|  |  | Neonates<sup>a</sup><br>(N=82) |  | 2-3-year-olds<sup>a</sup><br>(N=121) |  | 6-7-year-olds<sup>a</sup><br>(N=139) |  |
| --- | --- | --- | --- | --- | --- | --- | --- |
|  |  | PAE effect | *p* | PAE effect | *p* | PAE effect | *p* |
| Superior longitudinal fasciculus | L | - | ns | ↑ FA<br>↓ MD | 0.038<br>0.014 | - | ns |
|  | R | - | ns | ↑ FA<br>↓ MD | 0.041<br>0.048 | - | ns |
| Sagittal stratum | L | - | ns | ↑ FA<br>↓ MD | 0.017<br>0.015 | ↑ FA | 0.019 |
|  | R | - | ns | ↑ FA | 0.048 | - | ns |
| Inferior cerebellar peduncle | R | - | ns | - | ns | ↑ FA | 0.0197 |
| Superior cerebellar peduncle | L | - | ns | ↑ FA | 0.00015* | ↑ MD | 0.045 |
|  | R | - | ns | ↑ FA | 0.00153* | - | ns |

Abbreviations: PAE, prenatal alcohol exposure; FA, fractional anisotropy; MD, mean diffusivity; L/R, left or right hemisphere; ns, not significant.
<sup>a</sup> Model adjusting for sex and maternal education.
\* Survived FDR correction for multiple comparisons.

### 3.7. Post-hoc sensitivity analysis with prenatal tobacco exposure

We tested the robustness of our longitudinal trajectory findings for FA in the SCP and SLF by additionally adjusting for prenatal tobacco exposure (PTE). A chi-square test comparing nested models that included PTE, retained the significance of the PAE-by-time effect for FA in the SCP-L (p=0.002), corrected for multiple comparisons. The (uncorrected) PAE-by-time effect in the SLF-R (p=0.042) was also retained (Supplementary Table 5).

## 4. DISCUSSION

Our data suggest that effects of PAE influence white matter maturational processes throughout early childhood, a critical window for neurodevelopment. Both the PAE and control groups demonstrated significant increases in FA and decreases in MD in all tracts from the neonatal period to age 6-7 years, consistent with previous longitudinal studies of DTI during this period (Treit et al., 2013). However, children with PAE showed differential rates of FA and MD change in specific tracts, including the SCP-L and SLF-R, compared with controls over the study period. Children with PAE demonstrated generally higher FA in these tracts at age 2-3 years. These changes, measured across three time points in children in a birth cohort study in South Africa, suggest that PAE impacts white matter microstructure persistently across the first 6-7 years of life. This is the first study that, being of sufficient duration, has reported an association between PAE and white matter from birth to school age in the same cohort.

Incorporating early-life imaging of children into a longitudinal study has allowed us to identify specific time points when maximum PAE-related white matter changes are detectable in the early years. As evidenced by significant PAE-by-time effects over the three time points, this study has found that in specific regions there was a difference in the trajectory of change in white matter maturation over the pre-school years. Both the SCP-L and the SLF-R demonstrated altered trajectory patterns in FA and MD and in the SCR-R in MD for children with PAE compared to controls. Imaging data captured at critical time points over this 6-7-year period allowed us to observe relative change across early childhood through post-hoc pairwise comparisons.

On examination of pairwise time points, when including only the neonatal time point and the 6-7-year time point (without including the middle, 2-3-year time point), none of the tracts in our model demonstrated a difference in FA trajectory, though PAE-by-time effects for MD in the SLF-R, SCP-L, and SCR-R were evident. Cross-sectional differences at the neonatal and 6-7-year time points were minimal compared to the 2-3-year time point—children with PAE showed greater FA in the SS-L and ICP-R at age 6-7 years, as well as increased MD in the SCP-L at age 6-7 years. However, none of these associations survived multiple comparisons correction.

Conversely, when looking at the neonatal to 2-3-year time points, children with PAE demonstrated a more rapid increase in FA maturation in the SCP-L/R and UF-R compared with controls. Moreover, during this developmental period, children with PAE demonstrated altered FA and/or MD maturation in the SLF-L/R, SS-L/R, and SCP-L/R, resulting in cross-sectional group differences for FA in all of these tracts at age 2-3 years. The maturation of white matter is most rapid during the first two years of postnatal life as reflected in the dramatic changes in metrics describing white matter microstructural change over this time (Gilmore et al., 2018; Girault et al., 2019). It is unsurprising therefore, that during this period of high-intensity change, diffusion imaging would be sensitive to a wide group of regions potentially impacted by PAE.

Likewise, when only the 2-3- and 6-7-year time points were included, children with PAE showed differential rates of FA and/or MD change in the SLF-L/R, SS-R, SCP-L (FA change survived multiple comparisons correction), CC-G, CC-S, fornix, and SCR-L. Group differences in all of these tracts except the CC-G, CC-S, SCR-L and fornix were evident at the 2-3-year time point, but only in the SCP-L at the 6-7-year time point. As such, we observed a tapering of group differences that occurred from birth to age 2-3 years during the developmental period of 2-3 to 6-7 years. In general, for the concerned tracts, children with PAE demonstrated a slower rate of FA maturation than controls from the second to third time points, while FA growth in the controls continued to increase at a more rapid pace. These altered patterns of FA maturation between children with PAE and controls across the first 6-7 years of life would not have been observed without intermediate imaging time points. As such, our findings highlight the value of studies investigating not only measures across early life time periods, but repeating through childhood and adolescence in order to better understand the effects of PAE on longitudinal patterns of brain growth and maturation.

Key regions which appear to have the most robust and persistent trajectory changes across this period were the SLF and SCP bilaterally. These regions have been well-described as being affected by PAE (Fan et al., 2016; Lebel et al., 2008; Paolozza et al., 2017; Treit et al., 2013) and play important roles in neural networks serving language function, visuospatial functioning, motor regulation, learning, and memory(Lebel et al., 2012; Mattson et al., 2001). While this study is consistent with previous research in finding PAE trajectory effects in these important regions, it has also found a more marked rate of change in white matter development between PAE and unexposed children in early life (0 to 2-3 years), compared to the later pre-school years (2-3 to 6-7 years).

The SLF, present in each cerebral hemisphere, is one of the major white matter association tracts connecting parietal, occipital and temporal lobes with ipsilateral frontal cortices. It is known to play a role in networks serving language function, visuospatial functioning and working memory and to be critical for integrating information from multiple sensory modalities essential for complex cognitive processes. Lebel and colleagues (2008) have demonstrated that PAE is associated with altered development of the SLF during childhood and adolescence, with decreased FA in some segments of the SLF and increased FA in others. Our findings extend these previous studies into infancy, showing that the effects of PAE on SLF white matter maturation are seen by 2 years of age, are particularly impactful on the maturation trajectory during this early period, and have ongoing impact throughout childhood.

The SCP conveys selective afferents to the cerebellum and carries extensive efferent connections from all the cerebellar nuclei to the thalamus and other deep grey nuclei, serving important pathways for motor regulation, learning, and memory(Lebel et al., 2012; Mattson et al., 2001). The SCP is also involved in several cognitive functions, including attention, visuospatial abilities, and working memory. Previous research has suggested that PAE can affect the development of the SCP during early life, with alterations in microstructural properties persisting into adolescence (Lebel et al., 2012; Treit et al., 2013). This could have long-lasting effects on motor and cognitive functions that rely on cerebellar-thalamocortical pathways, such as motor coordination, learning, and memory. Separately, the SCR is one of the key projection tracts linking the brain stem with deep grey structures with the cerebral cortex, particularly the frontal and parietal lobes, supporting a wide range of cognitive, perceptual and motor systems.

Critically, the present study has succeeded in acquiring high quality neuroimaging in unsedated children, conventionally difficult at this age, and yet important, due to this being one of the most intense periods of brain development in terms of growth, maturation, and potential for neuroplasticity. However, the study was limited by its small sample size, which to some degree was related to participant dropout at follow-up time points, but also challenges with acquiring quality images in children as they are prone to move during scanning. Missing data is a challenging, yet inevitable aspect in most studies of longitudinal nature and with pediatric samples (Lebel et al., 2021). This also impacted our ability to test non-linear associations between PAE and white matter changes over time (Reynolds et al., 2019). Another potential limitation is the fact that maternal alcohol use during pregnancy was self-reported, which, due to social desirability bias and fear of stigma, may have resulted in under-reporting. Moreover, even though PAE status was confirmed in all cases, for this analysis we were not adequately powered to examine effects of the dosage and frequency of PAE on trajectories of white matter maturation. The impact of these factors, along with other potentially confounding pre-or postnatal exposures which we did not measure or account for, on associations between PAE and white matter maturation should be explored in future studies. Whether children with PAE will continue to show altered patters of white matter microstructural development beyond age 6-7 years should be further explored.

Overall, we describe altered trajectories of white matter maturation between different developmental periods (i.e., neonatal age to age 2-3 versus age 2-3 to 6-7 years), influenced by PAE. Regions identified in this study as showing altered white matter development trajectories in children with PAE have important functional connections to cognitive and motor functions, as well as social and emotional behaviours. This not only emphasizes the importance of acquiring early in-between time points in longitudinal neuroimaging studies of PAE, but also suggests differential sensitive periods of neurodevelopment and vulnerability to environmental exposures, as well as potential optimal windows of early intervention in children with PAE.

## ABBREVIATIONS

PAE, prenatal alcohol exposure; FA, fractional anisotropy; MD, mean diffusivity; FDR, false discovery rate; MRI, magnetic resonance imaging; DWI, diffusion weighted imaging; DCHS, Drakenstein Child Health Study; FASD, fetal alcohol spectrum disorder; SLF, superior longitudinal fasciculus; SS, sagittal stratum; UF, uncinate fasciculus; ICP, inferior cerebellar peduncle; SCP, superior cerebellar peduncle; CC-G, corpus callosum genu; CC-S, corpus callosum splenium; SCR, superior corona radiata.

## Data Availability

All data produced in the present study are available upon reasonable request to the authors.

## ACKNOWLEDGEMENTS

We greatly thank all mothers and children who participated in this study. We thank the DCHS team, and the clinical and administrative staff of the Western Cape Government Department of Health at Paarl Hospital and at the clinics for their support of the study. We also thank the radiographers at the Cape Universities Brain Imaging Centre (CUBIC) for their assistance with acquiring scans, Stefan du Plessis and Jean-Paul Fouché for the provision of custom batching-scripts to implement the image preprocessing pipeline, as well as the Centre for High Performance Computing (CHPC), South Africa, for providing computational resources to this project. Finally, we thank our collaborators and students for their scientific inputs.

## CRediT AUTHORSHIP CONTRIBUTION STATEMENT

**Kirsten Donald:** Conceptualization, Methodology, Investigation, Formal analysis, Writing – original draft, Supervision, Project administration, Funding acquisition, Writing – review & editing. **Chanellé Hendrikse:** Software, Writing – original draft, Data curation, Writing – review & editing. **Annerine Roos:** Software, Investigation, Data curation, Writing – review & editing. **Catherine Wedderburn:** Software, Investigation, Data curation, Writing – review & editing. **Sivenesi Subramoney:** Investigation, Writing – review & editing. **Jessica Ringshaw:** Investigation, Data curation, Writing – review & editing. **Layla Bradford:** Investigation, Data curation, Writing – review & editing. **Nadia Hoffman:** Project administration, Data curation, Writing – review & editing. **Tiffany Burd:** Project administration, Data curation, Writing – review & editing. **Katherine Narr:** Conceptualization, Methodology, Writing – review & editing. **Roger Woods:** Conceptualization, Methodology, Writing – review & editing. **Heather Zar:** Conceptualization, Methodology, Supervision, Funding acquisition, Writing – review & editing. **Shantanu Joshi:** Conceptualization, Methodology, Software, Formal analysis, Writing – original draft, Visualization, Writing – review & editing. **Dan Stein:** Conceptualization, Methodology, Supervision, Writing – review & editing.

## DATA AVAILABILITY

Study data is available from the corresponding author upon reasonable request.

## SUPPLEMENTARY MATERIAL

**Supplementary figure 1:**
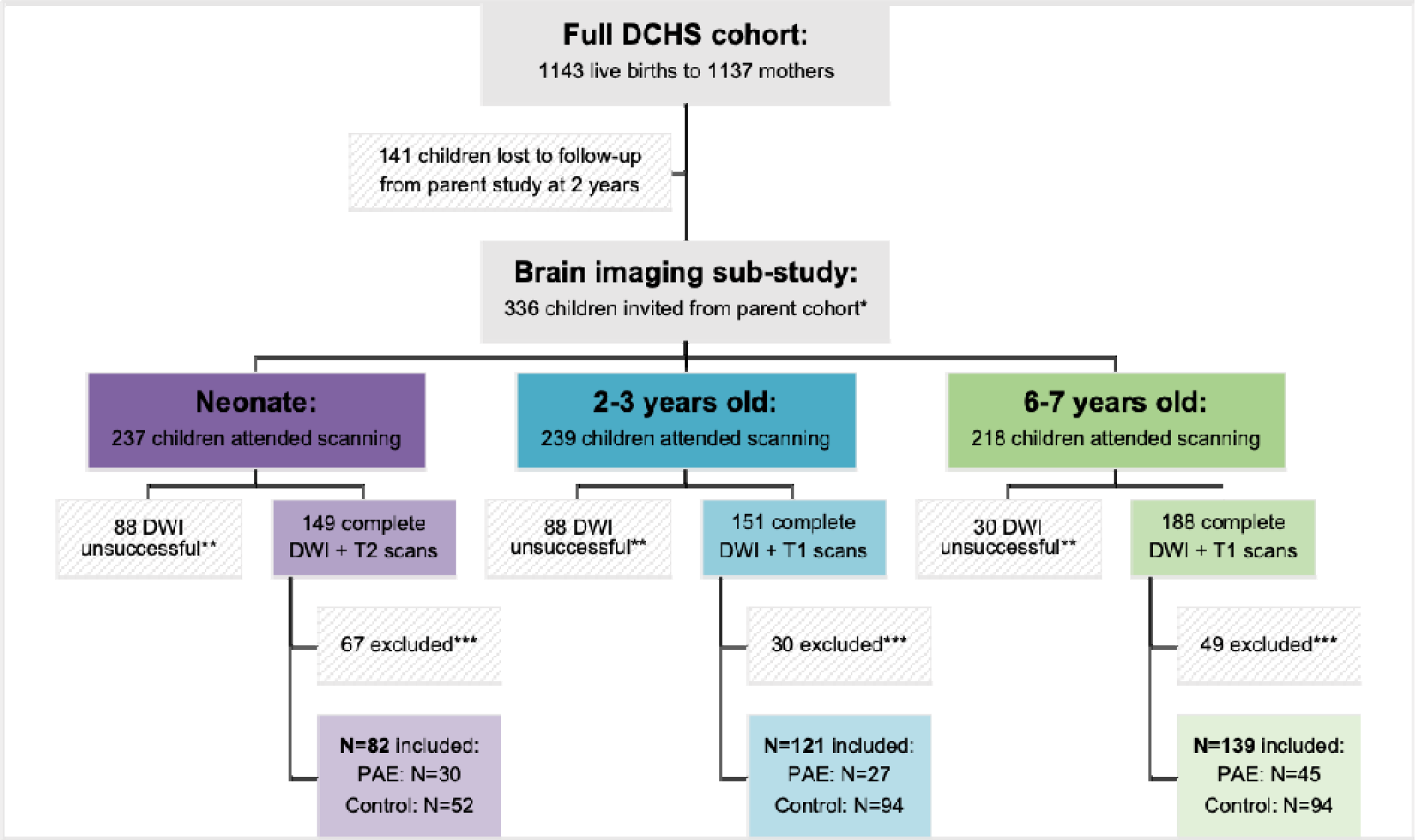
Flowchart of participation in the main Drakenstein Child Health Study (DCHS) and the current neuroimaging sub-study. *Exclusion criteria provided in main text. **Reasons for unsuccessful scans were: the child did not want to be scanned or was unable to sleep (neonatal and 2–3-year time points) during the scan, or excessive movement during scanning leading to premature termination of scan. ***Reasons for pre-or post-processing exclusion were: processing errors, or poor scan or output quality due to motion or other artifacts.

**Supplementary table 1:** Longitudinal effects of PAE, Time, and PAE-by-Time interaction on FA and MD.

| Tract | L/R | Fractional Anisotropy (FA)<sup>a</sup> |  |  |  |  |  | Mean diffusivity (MD)<sup>a</sup> |  |  |  |  |  |
| --- | --- | --- | --- | --- | --- | --- | --- | --- | --- | --- | --- | --- | --- |
|  |  | PAE |  | Time |  | PAE x Time |  | PAE |  | Time |  | PAE x Time |  |
|  |  | Partial eta<sup>2</sup> | *p* | Partial eta<sup>2</sup> | *p* | Partial eta<sup>2</sup> | *p* | Partial eta<sup>2</sup> | *p* | Partial eta<sup>2</sup> | *p* | Partial eta<sup>2</sup> | *p* |
| Superior longitudinal fasciculus | L | 0.016 | 0.025 | 0.842 | <0.001* | 0.002 | 0.733 | 0.001 | 0.656 | 0.962 | <0.001* | 0.019 | 0.055 |
|  | R | 0.009 | 0.143 | 0.864 | <0.001* | 0.027 | 0.046 | 0.016 | 0.031 | 0.951 | <0.001* | 0.022 | 0.038 |
| Sagittal stratum | L | 0.039 | 0.001 | 0.881 | <0.001* | 0.008 | 0.339 | 0.009 | 0.112 | 0.928 | <0.001* | 0.004 | 0.537 |
|  | R | 0.010 | 0.125 | 0.897 | <0.001* | 0.010 | 0.305 | 0.003 | 0.371 | 0.957 | <0.001* | 0.014 | 0.110 |
| Uncinate fasciculus | L | 0.000 | 0.985 | 0.923 | <0.001* | 0.011 | 0.235 | 0.001 | 0.618 | 0.935 | <0.001* | 0.004 | 0.505 |
|  | R | 0.002 | 0.442 | 0.820 | <0.001* | 0.017 | 0.080 | 0.000 | 0.821 | 0.690 | <0.001* | 0.008 | 0.303 |
| Inferior cerebellar peduncle | L | 0.012 | 0.075 | 0.563 | <0.001* | 0.005 | 0.554 | 0.001 | 0.608 | 0.744 | <0.001* | 0.001 | 0.886 |
|  | R | 0.026 | 0.011 | 0.512 | <0.001* | 0.003 | 0.728 | 0.000 | 0.828 | 0.262 | <0.001* | 0.001 | 0.871 |
| Superior cerebellar peduncle | L | 0.023 | 0.007 | 0.522 | <0.001* | 0.042 | 0.001* | 0.008 | 0.126 | 0.109 | <0.001* | 0.035 | 0.008 |
|  | R | 0.025 | 0.021 | 0.784 | <0.001* | 0.019 | 0.125 | 0.016 | 0.108 | 0.477 | <0.001* | 0.013 | 0.373 |
| Corpus callosum genu | n/a | 0.010 | 0.101 | 0.954 | <0.001* | 0.012 | 0.193 | 0.002 | 0.383 | 0.951 | <0.001* | 0.011 | 0.175 |
| Corpus callosum splenium | n/a | 0.007 | 0.225 | 0.924 | <0.001* | 0.001 | 0.876 | 0.001 | 0.607 | 0.978 | <0.001* | 0.008 | 0.408 |
| Fornix | n/a | 0.002 | 0.582 | 0.248 | <0.001* | 0.011 | 0.371 | 0.007 | 0.262 | 0.037 | 0.039 | 0.012 | 0.364 |
| Superior corona radiata | L | 0.005 | 0.248 | 0.926 | <0.001* | 0.016 | 0.131 | 0.008 | 0.136 | 0.968 | <0.001* | 0.019 | 0.059 |
|  | R | 0.010 | 0.118 | 0.924 | <0.001* | 0.003 | 0.670 | 0.007 | 0.146 | 0.965 | <0.001* | 0.021 | 0.047 |

Abbreviations: PAE, prenatal alcohol exposure; L/R, left or right hemisphere; n/a, not applicable.
<sup>a</sup> Model adjusting for sex and maternal education.
\* Survived FDR correction for multiple comparisons.

**Supplementary table 2:** Effects of PAE, Time, and PAE-by-time interaction on FA and MD (neonates and 2-3-year-olds).

| Tract | L/R | Fractional Anisotropy (FA)<sup>a</sup> |  |  |  |  |  | Mean diffusivity (MD)<sup>a</sup> |  |  |  |  |  |
| --- | --- | --- | --- | --- | --- | --- | --- | --- | --- | --- | --- | --- | --- |
|  |  | PAE |  | Time |  | PAE x Time |  | PAE |  | Time |  | PAE x Time |  |
|  |  | Partial eta<sup>2</sup> | *p* | Partial eta<sup>2</sup> | *p* | Partial eta<sup>2</sup> | *p* | Partial eta<sup>2</sup> | *p* | Partial eta<sup>2</sup> | *p* | Partial eta<sup>2</sup> | *p* |
| Superior longitudinal fasciculus | L | 0.055 | 0.007 | 0.372 | <0.001* | 0.003 | 0.531 | 0.006 | 0.265 | 0.917 | <0.001* | 0.004 | 0.372 |
|  | R | 0.036 | 0.015 | 0.818 | <0.001* | 0.000 | 0.929 | 0.025 | 0.027 | 0.929 | <0.001* | 0.006 | 0.283 |
| Sagittal stratum | L | 0.040 | 0.011 | 0.697 | <0.001* | 0.006 | 0.338 | 0.014 | 0.120 | 0.937 | <0.001* | 0.000 | 0.800 |
|  | R | 0.022 | 0.037 | 0.798 | <0.001* | 0.002 | 0.507 | 0.007 | 0.242 | 0.934 | <0.001* | 0.004 | 0.390 |
| Uncinate fasciculus | L | 0.004 | 0.425 | 0.474 | <0.001* | 0.002 | 0.540 | 0.005 | 0.367 | 0.894 | <0.001* | 0.002 | 0.508 |
|  | R | 0.001 | 0.744 | 0.815 | <0.001* | 0.026 | 0.032 | 0.000 | 0.895 | 0.429 | <0.001* | 0.013 | 0.145 |
| Inferior cerebellar peduncle | L | 0.011 | 0.245 | 0.574 | <0.001* | 0.002 | 0.586 | 0.000 | 0.804 | 0.812 | <0.001* | 0.000 | 0.840 |
|  | R | 0.011 | 0.134 | 0.322 | <0.001* | 0.000 | 0.994 | 0.000 | 0.835 | 0.205 | <0.001* | 0.003 | 0.529 |
| Superior cerebellar peduncle | L | 0.029 | 0.019 | 0.252 | <0.001* | 0.043 | 0.004 | 0.028 | 0.039 | 0.131 | <0.001* | 0.006 | 0.341 |
|  | R | 0.025 | 0.026 | 0.400 | <0.001* | 0.021 | 0.039 | 0.028 | 0.126 | 0.530 | <0.001* | 0.017 | 0.230 |
| Corpus callosum genu | n/a | 0.022 | 0.062 | 0.970 | <0.001* | 0.007 | 0.302 | 0.006 | 0.274 | 0.934 | <0.001* | 0.001 | 0.594 |
| Corpus callosum splenium | n/a | 0.010 | 0.187 | 0.838 | <0.001* | 0.001 | 0.714 | 0.005 | 0.352 | 0.964 | <0.001* | 0.002 | 0.552 |
| Fornix | n/a | 0.006 | 0.291 | 0.036 | 0.007 | 0.007 | 0.231 | 0.007 | 0.401 | 0.067 | 0.008 | 0.018 | 0.169 |
| Superior corona radiata | L | 0.018 | 0.057 | 0.922 | <0.001* | 0.000 | 0.996 | 0.016 | 0.078 | 0.959 | <0.001* | 0.006 | 0.291 |
|  | R | 0.016 | 0.109 | 0.886 | <0.001* | 0.000 | 0.884 | 0.014 | 0.101 | 0.946 | <0.001* | 0.007 | 0.261 |

Abbreviations: PAE, prenatal alcohol exposure; L/R, left or right hemisphere; n/a, not applicable.
<sup>a</sup> Model adjusting for sex and maternal education.
\* Survived FDR correction for multiple comparisons.

**Supplementary table 3:** Effects of PAE, Time, and PAE-by-time interaction on FA and MD (neonates and 6-7-year-olds).

| Tract | L/R | Fractional Anisotropy (FA)<sup>a</sup> |  |  |  |  |  | Mean diffusivity (MD)<sup>a</sup> |  |  |  |  |  |
| --- | --- | --- | --- | --- | --- | --- | --- | --- | --- | --- | --- | --- | --- |
|  |  | PAE |  | Time |  | PAE x Time |  | PAE |  | Time |  | PAE x Time |  |
|  |  | Partial eta<sup>2</sup> | *p* | Partial eta<sup>2</sup> | *p* | Partial eta<sup>2</sup> | *p* | Partial eta<sup>2</sup> | *p* | Partial eta<sup>2</sup> | *p* | Partial eta<sup>2</sup> | *p* |
| Superior longitudinal fasciculus | L | 0.014 | 0.086 | 0.822 | <0.001* | 0.002 | 0.530 | 0.003 | 0.432 | 0.969 | <0.001* | 0.005 | 0.357 |
|  | R | 0.004 | 0.430 | 0.895 | <0.001* | 0.021 | 0.078 | 0.011 | 0.118 | 0.944 | <0.001* | 0.022 | 0.028 |
| Sagittal stratum | L | 0.033 | 0.021 | 0.727 | <0.001* | 0.000 | 0.830 | 0.003 | 0.449 | 0.897 | <0.001* | 0.002 | 0.538 |
|  | R | 0.002 | 0.547 | 0.886 | <0.001* | 0.003 | 0.437 | 0.002 | 0.573 | 0.956 | <0.001* | 0.015 | 0.080 |
| Uncinate fasciculus | L | 0.002 | 0.571 | 0.871 | <0.001* | 0.004 | 0.425 | 0.000 | 0.790 | 0.949 | <0.001* | 0.005 | 0.278 |
|  | R | 0.000 | 0.790 | 0.862 | <0.001* | 0.016 | 0.066 | 0.001 | 0.628 | 0.822 | <0.001* | 0.009 | 0.221 |
| Inferior cerebellar peduncle | L | 0.012 | 0.192 | 0.703 | <0.001* | 0.011 | 0.207 | 0.002 | 0.599 | 0.124 | <0.001* | 0.000 | 0.807 |
|  | R | 0.021 | 0.033 | 0.491 | <0.001* | 0.002 | 0.531 | 0.002 | 0.587 | 0.256 | <0.001* | 0.003 | 0.486 |
| Superior cerebellar peduncle | L | 0.000 | 0.896 | 0.631 | <0.001* | 0.002 | 0.524 | 0.000 | 0.967 | 0.050 | <0.001* | 0.030 | 0.010 |
|  | R | 0.006 | 0.354 | 0.826 | <0.001* | 0.005 | 0.388 | 0.006 | 0.397 | 0.155 | <0.001* | 0.002 | 0.620 |
| Corpus callosum genu | n/a | 0.002 | 0.526 | 0.884 | <0.001* | 0.001 | 0.601 | 0.001 | 0.662 | 0.954 | <0.001* | 0.012 | 0.114 |
| Corpus callosum splenium | n/a | 0.011 | 0.237 | 0.940 | <0.001* | 0.001 | 0.680 | 0.000 | 0.788 | 0.973 | <0.001* | 0.003 | 0.471 |
| Fornix | n/a | 0.001 | 0.773 | 0.242 | <0.001* | 0.000 | 0.973 | 0.001 | 0.694 | 0.010 | 0.212 | 0.004 | 0.446 |
| Superior corona radiata | L | 0.001 | 0.690 | 0.923 | <0.001* | 0.010 | 0.165 | 0.016 | 0.078 | 0.959 | <0.001* | 0.006 | 0.291 |
|  | R | 0.008 | 0.246 | 0.937 | <0.001* | 0.003 | 0.486 | 0.005 | 0.292 | 0.963 | <0.001* | 0.022 | 0.033 |

Abbreviations: PAE, prenatal alcohol exposure; L/R, left or right hemisphere; n/a, not applicable.
<sup>a</sup> Model adjusting for sex and maternal education.
\* Survived FDR correction for multiple comparisons.

**Supplementary table 4:** Effects of PAE, Time, and PAE-by-time interaction on FA and MD (2-3-year-olds and 6-7-year-olds).

| Tract | L/R | Fractional Anisotropy (FA)<sup>a</sup> |  |  |  |  |  | Mean diffusivity (MD)<sup>a</sup> |  |  |  |  |  |
| --- | --- | --- | --- | --- | --- | --- | --- | --- | --- | --- | --- | --- | --- |
|  |  | PAE |  | Time |  | PAE x Time |  | PAE |  | Time |  | PAE x Time |  |
|  |  | Partial eta<sup>2</sup> | *p* | Partial eta<sup>2</sup> | *p* | Partial eta<sup>2</sup> | *p* | Partial eta<sup>2</sup> | *p* | Partial eta<sup>2</sup> | *p* | Partial eta<sup>2</sup> | *p* |
| Superior longitudinal fasciculus | L | 0.010 | 0.110 | 0.778 | <0.001* | 0.000 | 0.875 | 0.003 | 0.477 | 0.909 | <0.001* | 0.072 | 0.001* |
|  | R | 0.005 | 0.435 | 0.155 | <0.001* | 0.051 | 0.008 | 0.009 | 0.290 | 0.473 | <0.001* | 0.039 | 0.028 |
| Sagittal stratum | L | 0.054 | <0.001* | 0.907 | <0.001* | 0.009 | 0.171 | 0.015 | 0.102 | 0.538 | <0.001* | 0.009 | 0.199 |
|  | R | 0.013 | 0.177 | 0.544 | <0.001* | 0.022 | 0.086 | 0.000 | 0.913 | 0.803 | <0.001* | 0.057 | 0.010* |
| Uncinate fasciculus | L | 0.000 | 0.913 | 0.938 | <0.001* | 0.013 | 0.103 | 0.000 | 0.877 | 0.674 | <0.001* | 0.002 | 0.513 |
|  | R | 0.015 | 0.083 | 0.024 | <0.001* | 0.001 | 0.663 | 0.005 | 0.303 | 0.374 | <0.001* | 0.002 | 0.525 |
| Inferior cerebellar peduncle | L | 0.021 | 0.042 | 0.101 | <0.001* | 0.001 | 0.678 | 0.001 | 0.682 | 0.783 | <0.001* | 0.002 | 0.540 |
|  | R | 0.048 | 0.010* | 0.139 | <0.001* | 0.002 | 0.645 | 0.002 | 0.654 | 0.034 | 0.049 | 0.002 | 0.647 |
| Superior cerebellar peduncle | L | 0.070 | <0.001* | 0.304 | <0.001* | 0.052 | <0.001* | 0.006 | 0.257 | 0.067 | <0.001* | 0.038 | 0.005* |
|  | R | 0.072 | 0.003* | 0.692 | <0.001* | 0.025 | 0.095 | 0.029 | 0.107 | 0.454 | <0.001* | 0.017 | 0.222 |
| Corpus callosum genu | n/a | 0.015 | 0.129 | 0.931 | <0.001* | 0.026 | 0.048 | 0.000 | 0.842 | 0.132 | <0.001* | 0.029 | 0.030 |
| Corpus callosum splenium | n/a | 0.010 | 0.311 | 0.593 | <0.001* | 0.010 | 0.301 | 0.001 | 0.727 | 0.649 | <0.001* | 0.042 | 0.041 |
| Fornix | n/a | 0.008 | 0.388 | 0.154 | <0.001* | 0.057 | 0.022 | 0.022 | 0.132 | 0.027 | 0.096 | 0.005 | 0.498 |
| Superior corona radiata | L | 0.004 | 0.444 | 0.251 | <0.001* | 0.043 | 0.017 | 0.001 | 0.674 | 0.312 | <0.001* | 0.032 | 0.042 |
|  | R | 0.010 | 0.230 | 0.408 | <0.001* | 0.004 | 0.450 | 0.001 | 0.802 | 0.750 | <0.001* | 0.026 | 0.090 |

Abbreviations: PAE, prenatal alcohol exposure; L/R, left or right hemisphere; n/a, not applicable.
<sup>a</sup> Model adjusting for sex and maternal education.
\* Survived FDR correction for multiple comparisons.

**Supplementary table 5:** Longitudinal effects of PAE, Time and PAE-by-Time on FA with and without adjusting for prenatal tobacco exposure.

|  |  | Model without PTE<sup>a</sup> |  |  | Model with PTE<sup>b</sup> |  |  |
| --- | --- | --- | --- | --- | --- | --- | --- |
| Tract | L/R | PAE | Time | PAE x Time | PAE | Time | PAE x Time |
| Superior longitudinal fasciculus | L | 0.025 | <0.001* | ns | ns | <0.001* | ns |
|  | R | ns | <0.001* | 0.046 | ns | <0.001* | 0.042 |
| Superior cerebellar peduncle | L | 0.007 | <0.001* | 0.001* | ns | <0.001* | 0.002* |
|  | R | 0.021 | <0.001* | 0.125 | ns | <0.001* | ns |

Abbreviations: PTE, prenatal tobacco exposure; PAE, prenatal alcohol exposure; FA, fractional anisotropy; L/R, left or right hemisphere; ns, not significant at p < 0.05.
<sup>a</sup> Model adjusting for sex and maternal education.
<sup>b</sup> Model adjusting for sex, maternal education, and prenatal tobacco exposure.
\* Survived FDR correction for multiple comparisons.

